# Clinical Profile of First 1000 COVID-19 Cases Admitted at Tertiary Care Hospitals and the Correlates of their Mortality: An Indian Experience

**DOI:** 10.1101/2020.11.16.20232223

**Authors:** Sandeep Budhiraja, Aakriti Soni, Vinitaa Jha, Abhaya Indrayan, Arun Dewan, Omender Singh, Yogendra Pal Singh, Indermohan Chugh, Vijay Arora, Rajesh Pande, Abdul Ansari, Sujeet Jha

## Abstract

**Objective:** To describe the clinical profile and factors leading to increased mortality in coronavirus disease (COVID-19) patients admitted to a group of hospitals in India.

**Design:** A records-based study of the first 1000 patients with a positive result on real-time reverse transcriptase-polymerase-chain-reaction assay for SARS-CoV-2 admitted to our facilities. Various factors such as demographics, presenting symptoms, co-morbidities, ICU admission, oxygen requirement and ventilator therapy were studied.

**Results:** Of the 1000 patients, 24 patients were excluded due to lack of sufficient data. Of the remaining 976 in the early phase of the epidemic, males were admitted twice as much as females (67.1% and 32.9%, respectively). Mortality in this initial phase was 10.6% and slightly higher for males and steeply higher for older patients. More than 8% reported no symptoms and the most common presenting symptoms were fever (78.3%), productive cough (37.2%), and dyspnea (30.64%). More than one-half (53.6%) had no co-morbidity. The major co-morbidities were hypertension (23.7%), diabetes without (15.4%), and with complications (9.6%). The co-morbidities were associated with higher ICU admissions, greater use of ventilators as well as higher mortality. A total of 29.9% were admitted to the ICU, with a mortality rate of 32.2%. Mortality was steeply higher in those requiring ventilator support (55.4%) versus those who never required ventilation (1.4%). The total duration of hospital stay was just a day longer in patients admitted to the ICU than those who remained in wards.

**Conclusion:** Male patients above the age of 60 and with co-morbidities faced the highest rates of mortality. They should be admitted to the hospital in early stage of the disease and given aggressive treatment to help reduce the morbidity and mortality associated with COVID-19.

## INTRODUCTION

An unprecedented disease hit the world some time ago. The World Health Organization (WHO) declared this a corona virus disease-19 (COVID-19) pandemic on 11^th^ March 2020. It has affected over 21,756,357 and killed 771,635 people by 19/08/2020^(1)^ when this report was prepared Many hospitals are now flooded with patients suffering from the COVID-19, presenting with a wide array of symptoms ranging from fever and respiratory distress to gastrointestinal symptoms. The death count been rapidly rising though the level (5.6%) has not reached the infliction by the other members of the coronavirus family causing human illnesses such SARS (13%) and MERS (35%)^(2)^.However, COVID is far more transmissible with an estimated reproduction number (R_0_) 3.32. ^(3)^

India too now is in the grip of the pandemic and, in terms of absolute numbers, the 3^rd^ worst affected country after USA and Brazil. Early countrywide lockdown helped in delaying the spread and shift the peak, and gave time to create infrastructure to face the surge. Despite this a total of 2,768,670 cases and 53,026 deaths have been reported in India as of 19/08/2020 ^(4)^. This enormous number has overwhelmed the medical system and has resulted in shortages of medical equipment and personal protective equipment (PPE) ^(5,6,7)^. Many hospitals across the country were designated as COVID-19 hospitals. Some hospitals in our group were recognized as official sites for managing COVID-19 patients on 27/03/2020 when the disease started to occur in the epidemic proportion in India. Multiple changes were made since then in our infrastructure to accommodate the large inflow of the cases.

Despite the swift spread and the rapidly increasing number of people getting affected, the complete clinical course of this disease is still unclear for Indian patients. Reports describing demographics, clinical characteristics, hospital course, morbidity, and mortality in patients in the Indian setting have been published ^(8,9,10)^ but they are based on limited numbers of cases.

We report a relatively large study of 1000 patients with known outcome to better understand the disease process and progression of COVID-19 cases, and to study the factors affecting the outcome. This may help in triaging the rapid rise of patients and streamlining resources for better management of cases with optimal efficiency and better outcomes.

## MATERIAL AND METHODS

### Study Design and Population

This is a multi-center, record based, observational study of patients admitted to 5 multi-specialty hospitals across Delhi and Mumbai that are designated as COVID-19 hospitals by the State Governments. All these are private hospitals and cater to generally well-to-do sections of the society. Over 7000 patients have been admitted to these hospitals till the middle of August 2020. We retrieved electronic and paper based medical records for the first 1000 hospitalized patients with laboratory confirmed COVID-19 diagnosis. These were admitted between 1^st^ April 2020 and 31^st^ May 2020.

The inclusion criteria were defined as:

1. Positive result on real-time reverse transcriptase–polymerase-chain-reaction (RT-PCR) assay of nasal and pharyngeal swab specimens for Severe Acute Respiratory Syndrome Coronavirus-2 (SARS-CoV-2).
2. Age > 18 years.
3. Closed cases with known outcome.

The Government testing guidelines changed twice during our study and has been mentioned in Box 1 for record. Although most of the mild cases now are being asked to home quarantine, at the time of this study, many with a positive RT-PCR were unable to quarantine and were admitted in the hospital. This the study includes mild as well as moderate and severe cases. The classification of cases into these different categories was based on the characteristics given in Box 2.

### Data Collection

The extracted data included basic demographics (age and gender), symptoms, co-morbidities, admission pathway (ward or ICU), treatment, transfer to ICU, oxygen therapy, ventilator requirement, duration of hospitalization, and the outcome. The study investigators checked the collected data independently. A separate dedicated quality control abstractor reviewed records with missing data or with inconsistent values and did corrections as much as possible.

The signs and symptoms included those seen in influenza-like illness (ILI)^(11)^, pneumonia (chest pain, dyspnea, wheezing, lower chest wall in drawing, history of TB), gastroenteritis (nausea, vomiting, abdominal pain, diarrhea), ear pain, altered consciousness, and seizures.

The co-morbidities included hypertension (HTN), diabetes with and without complications, obesity, chronic kidney disease (CKD), moderate or severe liver disease, asthma, chronic pulmonary disease other than asthma, and chronic cardiac disease including congenital anomalies but excluding HTN. We also analyzed the outcome based on the route of admission (ICU or ward), oxygen therapy and ventilator requirement.

#### Box 1

##### Criteria for coronavirus disease 2019 (COVID-19) testing and diagnosis

###### Testing policies

**Strategy for COVID19 testing in India**

**April (09/04/2020):**

1. All symptomatic individuals who have undertaken international travel in the last 14 days
2. All symptomatic contacts of laboratory confirmed cases
3. All symptomatic health care workers
4. All patients with Severe Acute Respiratory Illness (SARI)
5. Asymptomatic direct and high-risk contacts of a confirmed case should be tested once between day 5 and day 14 of coming in his/her contact
6. All symptomatic Influenza like illness (ILI)-fever, cough, sore throat, runny nose

**In hotspots/cluster and in large migration gatherings/ evacuees centres Mid May (18/05/2020):**

1. All symptomatic (ILI symptoms) individuals with history of international travel in the last 14 days.
2. All symptomatic (ILI symptoms) contacts of laboratory confirmed cases.
3. All symptomatic (ILI symptoms) health care workers / frontline workers involved in containment and mitigation of COVID19.
4. All patients of Severe Acute Respiratory Infection (SARI).
5. Asymptomatic direct and high-risk contacts of a confirmed case to be tested once between day 5 and day 10 of coming into contact.
6. All symptomatic ILI within hotspots/containment zones.
7. All hospitalised patients who develop ILI symptoms.
8. All symptomatic ILI among returnees and migrants within 7 days of illness.
9. No emergency procedure (including deliveries) should be delayed for lack of test. However, sample can be sent for testing if indicated as above (1-8), simultaneously.

*ILI case is defined as one with acute respiratory infection with fever* ≥ *38*°*C AND cough*.

*SARI case is defined as one with acute respiratory infection with fever* ≥ *38*°*C AND cough AND requiring hospitalization*.

###### Diagnosis

A covid-19 diagnosis was defined as a positive result on the reverse transcriptase polymerase chain reaction assay for severe acute respiratory syndrome coronavirus 2

#### Box 2

##### Stratification of patients

###### Mild cases

Fever, myalgia, cough, headache, sore throat, nasal congestion, fatigue, diarrhea, loss of smell/taste preceding respiratory symptoms.

###### Mild cases with risk factors

Age more than 60

Medical Comorbidities:

- CAD
- Diabetes
- HT
- Transplant
- Immunosuppression
- Cancer
- Kidney disease
- Obesity: BMI:25KG/M2

###### ≥ 2 of the following

- Any symptom of mild disease
- Dyspnea, respiratory rate more than or equal to 24/min
- Hypoxia (SPO2 less than equal to 94%: range 90-94%)
- No additional signs or symptoms of severe COVID 19
- X Ray chest or lung ultrasound with bilateral ground glass opacities or bilateral consolidation

###### Severe cases (including immunocompromised, HIV, transplant, malignancy patients): ≥ 2 of the following

- Respiratory rate>30/min; SPO2<90%
- Severe respiratory distress
- Clinical signs of pneumonia
- Require mechanical ventilation
- Not on pressors, creatinine clearance more 30 ml per min, ALT less than 5x upper limit of normal
- Radiographic imaging with bilateral ground glass opacities of consolidation
- ARDS with PaO2/FiO2 151 to 300mm Hg,
- Lymphopenia
- IL6 > 40 pg/ml or CRP > 10 mg/l or elevated d-dimer with no other suspected cause

###### ≥ 2 of the following

- Patient on mechanical ventilation
- Radiographic imaging with bilateral ground glass opacities of consolidation
- ARDS with PaO2/FiO2 less than equal to 150 mm Hg
- Septic Shock requiring pressors
- Altered consciousness
- Multi organ failure as evidenced by CrCL<30 ml/min or receiving HD, CVVH or ALT > 5x upper limit of normal.

### Statistical Analysis

The primary end point was death or discharged alive. The collected data was processed using different cross-tabulations to analyze which characteristics were highly linked to mortality. Chi-square test (χ^2^) and Fisher exact test (for small cell frequencies) were used for checking statistical significance of the association. Another chi-square test for linear trend was used to assess the statistical significance of the trend. Duration of hospital and ICU stay was evaluated in terms of median and interquartile range (IQR) because of their highly skewed distribution. Difference in these was assessed using Mann-Whitney-Wilcoxon test. The level of significance was 5%. SPSS 20.0 (IBM Corp., Armonk, NY, US) was used for all the calculations.

## RESULTS

### Demographics and Baseline

Out of the 1000 patients in the study, 24 were excluded due to incomplete data. Median age was 47.5 years (interquartile range: 34.5-59.9 years). Nearly one-fourth (24.7%) of the patients were of the age 60 years or more (Table 1) against around 10% population of this age in this area. Thus, disproportionately higher percentages of old-age patients were admitted with COVID-19.

**Table 1.** Age distribution of cases by sex, number of symptoms, and the number of co-morbidities, requirement of ventilator and ICU, and deaths.

| Characteristic | Age -Group (years) |  |  |  |  |  |  |  |  |  |
| --- | --- | --- | --- | --- | --- | --- | --- | --- | --- | --- |
|  | Total |  | <45 |  | 45-59 |  | 60-74 |  | ≥75 |  |
|  | Count | % | Count | % | Count | % | Count | % | Count | % |
| All Cases | 976 | 100.0 | 441 | 45.18 | 293 | 30.02 | 196 | 20.08 | 46 | 4.71 |
| **Sex** |  |  |  |  |  |  |  |  |  |  |
| Male | 655 | 67.11 | 294 | 66.67 | 192 | 65.53 | 136 | 69.39 | 33 | 71.74 |
| Female | 321 | 32.89 | 147 | 33.33 | 101 | 34.47 | 60 | 30.61 | 13 | 28.26 |
| **No. of symptoms** |  |  |  |  |  |  |  |  |  |  |
| 0 | 84 | 8.61 | 38 | 8.62 | 23 | 7.85 | 19 | 9.69 | 4 | 8.70 |
| 1 | 205 | 21.00 | 96 | 21.77 | 56 | 19.11 | 44 | 22.45 | 9 | 19.57 |
| 2 | 349 | 35.76 | 146 | 33.11 | 104 | 35.49 | 80 | 40.82 | 19 | 41.30 |
| 3 or more | 338 | 34.63 | 161 | 36.51 | 110 | 37.54 | 53 | 27.04 | 14 | 30.43 |
| **No. of comorbidities** |  |  |  |  |  |  |  |  |  |  |
| 0 | 523 | 53.59 | 355 | 80.50 | 113 | 38.57 | 43 | 21.94 | 12 | 26.09 |
| 1 | 178 | 18.24 | 49 | 11.11 | 81 | 27.65 | 42 | 21.43 | 6 | 13.04 |
| 2 | 156 | 15.98 | 26 | 5.90 | 67 | 22.87 | 53 | 27.04 | 10 | 21.74 |
| 3 or more | 119 | 12.19 | 11 | 2.49 | 32 | 10.92 | 58 | 29.59 | 18 | 39.13 |
| **ICU/Ventilator/Death** |  |  |  |  |  |  |  |  |  |  |
| ICU | 292 | 29.92 | 86 | 19.50 | 86 | 29.35 | 86 | 43.88 | 34 | 73.91 |
| Ventilator | 166 | 17.01 | 17 | 3.85 | 64 | 21.84 | 61 | 31.12 | 24 | 52.17 |
| Death | 103 | 10.55 | 8 | 1.81 | 29 | 9.90 | 47 | 23.98 | 19 | 41.30 |

Of these 976 cases, 655 (67.11%) were males and 321 (32.89%) were females. Males were significantly (P < 0.001) higher and the male-female ratio was nearly 2:1. In the age group 45-59 years, the ratio for males was slightly less but for the age group 75+ years the ratio for males was significantly higher than the overall average. If the admissions were in proportion of the cases in the community, our data suggests that the older age males were more commonly affected.

Details of the specific symptoms and co-morbidities are provided later. Of the total admissions, 8.61% were asymptomatic and this percentage remained nearly the same across all age groups. Nearly one-third (35.76%) reported two symptoms and another one-third (34.63%) reported more than 3 symptoms (Table 1). The number of symptoms was not associated with age (P = 0.469). More than half (53.59%) had no co-morbidity and 12.19% of the admitted patients had 3 or more co-morbidities (Table 1). Against only 2.49% with 3 or more co-morbidity in the age-group less than 45 years, more than 10 times (29.59%) of the age 75 years or more had 3 or more co-morbidities. This difference is statistically highly significant (P < 0.001). Nearly 30% had to be admitted to ICU and nearly 17% had to be on ventilator. Both significantly (P < 0.001) increased with the age of the patients.

### Correlates of Mortality

The main outcome under study is death or discharged alive. Of the 976 patients with a known outcome, 873 (89.45%) were discharged alive and 103 (10.5%) died in the hospital. Age is significantly associated with outcome (P < 0.001) in both males and females. Higher the age the more was the mortality. More importantly, mortality in patients 75 years and above was 4 times the mortality in age 45-59 in both the sexes (Table 2). Sex on the other hand as an individual characteristic failed to be significantly associated with mortality (P = 0.127) although mortality was slightly higher in males than females (11.60% vs. 8.41%).

**Table 2.**
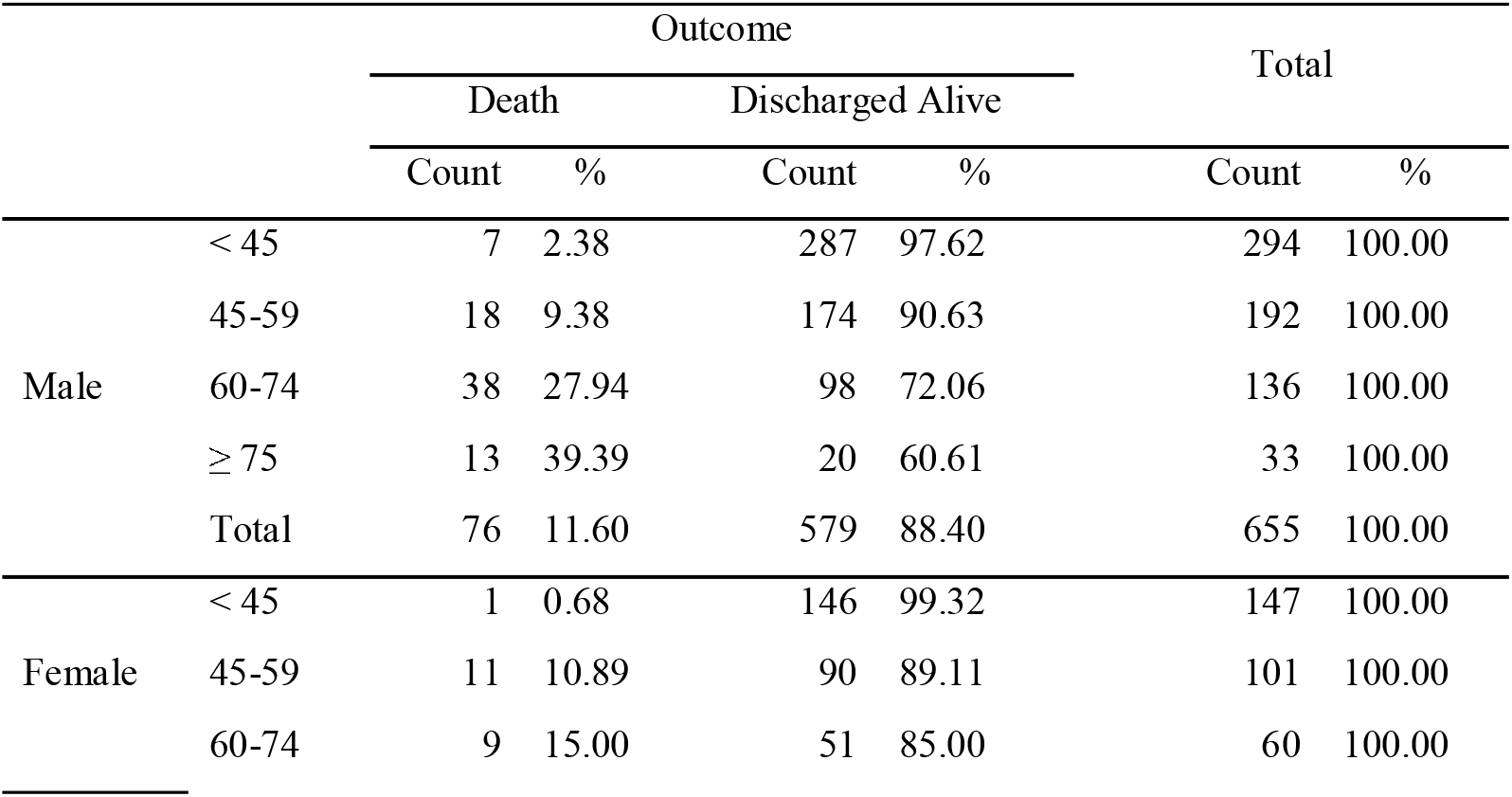
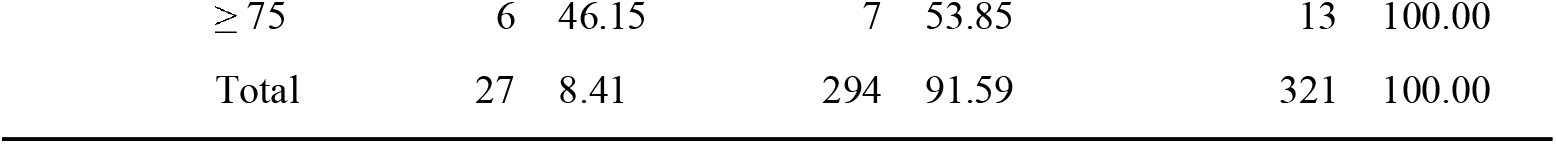
Mortality by age-group and sex.

The highest (46.15%) mortality was seen in females of age 75+, followed by males (39.39%) of the same age group. However, this difference in mortality between males and females in this age group is not statistically significant (P = 0.675). In the age-group 60-74 years, the mortality was 27.94% in males and 15.00% in females, and this difference is significant (P = 0.05).

More than one-half (53.58%) admitted patients reported no co-morbidity. The most common co-morbidities in admitted patients with COVID-19 were hypertension (23.67%), diabetes without macrovascular complications (15.37%), diabetes with macrovascular complications (9.63%), chronic cardiac disease (5.64%), and chronic kidney disease (5.43%) (Table 3). Less common co-morbidities were chronic pulmonary disease (7 cases), obesity (4 cases), chronic neurological disorder (3 cases), and liver disease (2 cases). Mortality was highest amongst those with chronic kidney disease (49.06), diabetes with complications (38.30%), followed by those with chronic cardiac disease (32.73%). Mortality steeply increased from 2.68% in those with no co-morbidity to 36.13% with 3 or more co-morbidities (P<0.001) (Table 4). Old age and high number of co-morbidities together substantially increased the rate of mortality (Figure 1). Requirement of admission to ICU (P < 0.001) and ventilator (P < 0.001) also steadily increased as the number of co-morbidities increased.

**Table 3.** Outcome in relation to the specific co-morbidities.

| Co-morbidity | Outcome |  |  |  | Total |  |
| --- | --- | --- | --- | --- | --- | --- |
|  | Death |  | Discharged Alive |  |  |  |
|  | Count | % | Count | % | Count | % |
| All Cases | 103 | 10.55 | 873 | 89.45 | 976 | 100.00 |
| Chronic cardiac disease, including congenital heart disease (Not hypertension) | 18 | 32.73 | 37 | 67.27 | 55 | 5.64 |
| Chronic kidney disease | 26 | 49.06 | 27 | 50.94 | 53 | 5.43 |
| Diabetes with macrovascular complications | 36 | 38.30 | 58 | 61.70 | 94 | 9.63 |
| Diabetes without macrovascular complications | 23 | 15.33 | 127 | 84.67 | 150 | 15.37 |
| Hypertension | 47 | 20.35 | 184 | 79.65 | 231 | 23.67 |

**Table 4.** Ventilator, ICU and outcomes in relation to co-morbidities.

| Number of Co-Morbidities | Use of Ventilator |  | Admission to ICU |  | Outcome |  |  |  | Total |  |
| --- | --- | --- | --- | --- | --- | --- | --- | --- | --- | --- |
|  |  |  |  |  | Death |  | Discharged Alive |  |  |  |
|  | Count | % | Count | % | Count | % | Count | % | Count | % |
| All Cases | 166 | 17.0 | 292 | 29.92 | 103 | 10.55 | 873 | 89.45 | 976 | 100.00 |
| 0 | 29 | 5.5 | 104 | 19.89 | 14 | 2.68 | 509 | 97.32 | 523 | 53.58 |
| 1 | 32 | 18.0 | 45 | 25.28 | 20 | 11.24 | 158 | 88.76 | 178 | 18.24 |
| 2 | 46 | 29.5 | 68 | 43.59 | 26 | 16.67 | 130 | 83.33 | 156 | 15.98 |
| 3 or more | 59 | 49.6 | 75 | 63.03 | 43 | 36.13 | 76 | 63.87 | 119 | 12.19 |

**Figure 1.**
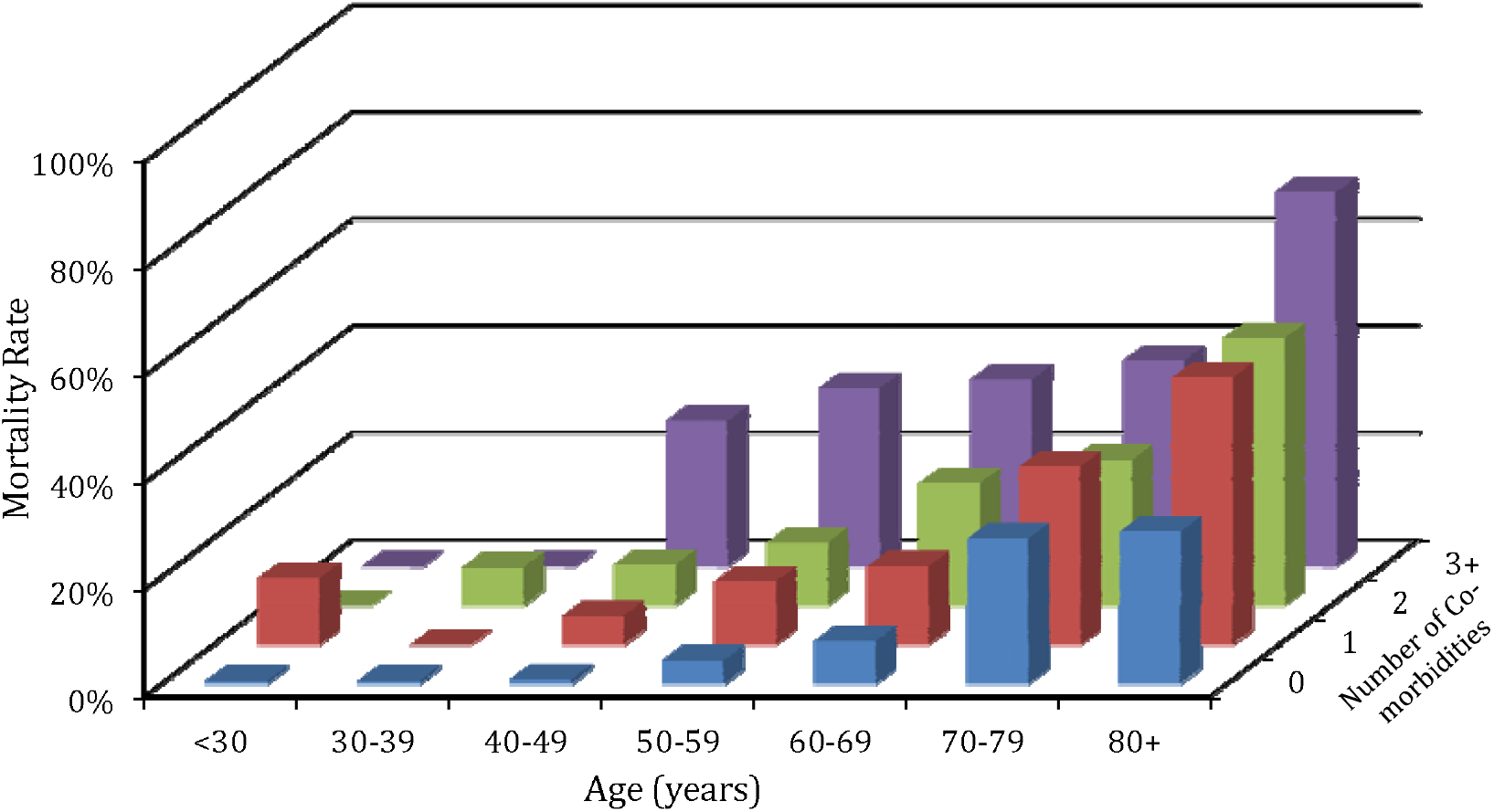
Mortality in relation to the age-group and the number of co-morbidities. Less than one-tenth (8.61%) of the admitted patients reported no symptom. The most common presenting symptoms were fever (78.28%), productive cough (37.19%), dyspnea (30.64%), and sore throat (25.9%). Mortality was disproportionately higher (24.41%) in those who were admitted with the complaint of dyspnea compared to the overall average. Cases with sore throat had less (3.95%) mortality than the overall average (Table 5). Mortality increased with the number of symptoms (Table 6) but the trend was not statistically significant (P = 0.072).

An increase in the number of symptoms resulted in increased chances of ICU admission and need for ventilator support. It also resulted in more deaths (Table 6). A similar trend was observed with increasing number of co-morbidities with mortality. However, the difference was statistically significant (P < 0.001) only for the number of co-morbidities, pointing to the increased risk of ICU admission, ventilator requirement and eventual death in patients with increasing co-morbidities and not with the number of symptoms.

**Table 5.** Outcomes in relation to common symptoms.

| Symptoms | Outcome |  |  |  | Total |  |
| --- | --- | --- | --- | --- | --- | --- |
|  | Death |  | Discharged Alive |  |  |  |
|  | Count | % | Count | % | Count | % |
| All Cases | 103 | 10.55 | 873 | 89.45 | 976 | 100.00 |
| History of fever | 75 | 9.82 | 689 | 90.18 | 764 | 78.28 |
| Cough with sputum production<br>bloody sputum/hemoptysis | 32 | 8.82 | 331 | 91.18 | 363 | 37.19 |
| Sore throat | 10 | 3.95 | 243 | 96.05 | 253 | 25.92 |
| Shortness of breath (Dyspnea) | 73 | 24.41 | 226 | 75.59 | 299 | 30.64 |

**Table 6.** Requirement of ventilator, ICU admission and the outcome depending upon the number of symptoms and co-morbidities.

| Number of Symptoms | Use of Ventilator |  | Admission to ICU |  | Outcome |  |  |  | Total |  |
| --- | --- | --- | --- | --- | --- | --- | --- | --- | --- | --- |
|  |  |  |  |  | Death |  | Discharged Alive |  |  |  |
|  | Count | % | Count | % | Count | % | Count | % | Count | % |
| All Cases | 166 | 17.0 | 292 | 29.92 | 103 | 10.55 | 873 | 89.45 | 976 | 100.00 |
| 0 | 6 | 7.1 | 16 | 19.05 | 7 | 8.33 | 77 | 91.67 | 84 | 8.61 |
| 1 | 32 | 15.6 | 51 | 24.88 | 20 | 9.76 | 185 | 90.24 | 205 | 21.00 |
| 2 | 62 | 17.8 | 103 | 29.51 | 35 | 10.03 | 314 | 89.97 | 349 | 35.76 |
| 3 or more | 66 | 19.5 | 122 | 36.09 | 41 | 12.13 | 297 | 87.87 | 338 | 34.63 |

### Hospital Course

Of the 976 patients, 292 (29.92%) were admitted to the ICU at least once during their stay in the hospital – 184 (63.01%) of them were directly admitted to the ICU and 108 (36.99%) had to be shifted from ward to ICU (Figure 2). Mortality is significantly (P < 0.001) and steeply higher (31.84%) in patients who are admitted to the ICU compared to the mortality (1.46%) in those not admitted to ICU (Table 7). However, no such significant (P = 0.949) difference in mortality is observed between those directly admitted to ICU (58/184) versus those who were shifted from ward to the ICU (35/108). In both these cases, the mortality was nearly 32%.

**Table 7.** Outcome in relation to the hospital course of the patients.

| Hospital Course | Outcome |  |  |  | Total |  |
| --- | --- | --- | --- | --- | --- | --- |
|  | Death |  | Discharged Alive |  |  |  |
|  | Count | % | Count | % | Count | % |
| All Cases | 103 | 10.55 | 873 | 89.45 | 976 | 100.00 |
| **Admission to ICU** |  |  |  |  |  |  |
| No ICU | 10 | 1.46 | 674 | 98.54 | 684 | 70.08 |
| Direct ICU | 58 | 31.52 | 126 | 68.48 | 184 | 18.85 |
| Ward to ICU | 35 | 32.41 | 73 | 67.59 | 108 | 11.07 |
| **Ventilator** |  |  |  |  |  |  |
| No | 11 | 1.36 | 799 | 98.64 | 810 | 82.99 |
| Yes | 92 | 55.42 | 74 | 44.58 | 166 | 17.01 |
| **Type of Ventilator** |  |  |  |  |  |  |
| No Ventilator | 11 | 1.36 | 799 | 98.64 | 810 | 82.99 |
| Invasive Ventilator | 47 | 82.46 | 10 | 17.54 | 57 | 5.84 |
| Non-Invasive Ventilator | 45 | 41.28 | 64 | 58.71 | 109 | 11.16 |
| **Oxygen Therapy** |  |  |  |  |  |  |
| No | 24 | 3.23 | 719 | 96.77 | 743 | 76.13 |
| Yes | 79 | 33.91 | 154 | 66.09 | 233 | 23.87 |

**Figure 2:**
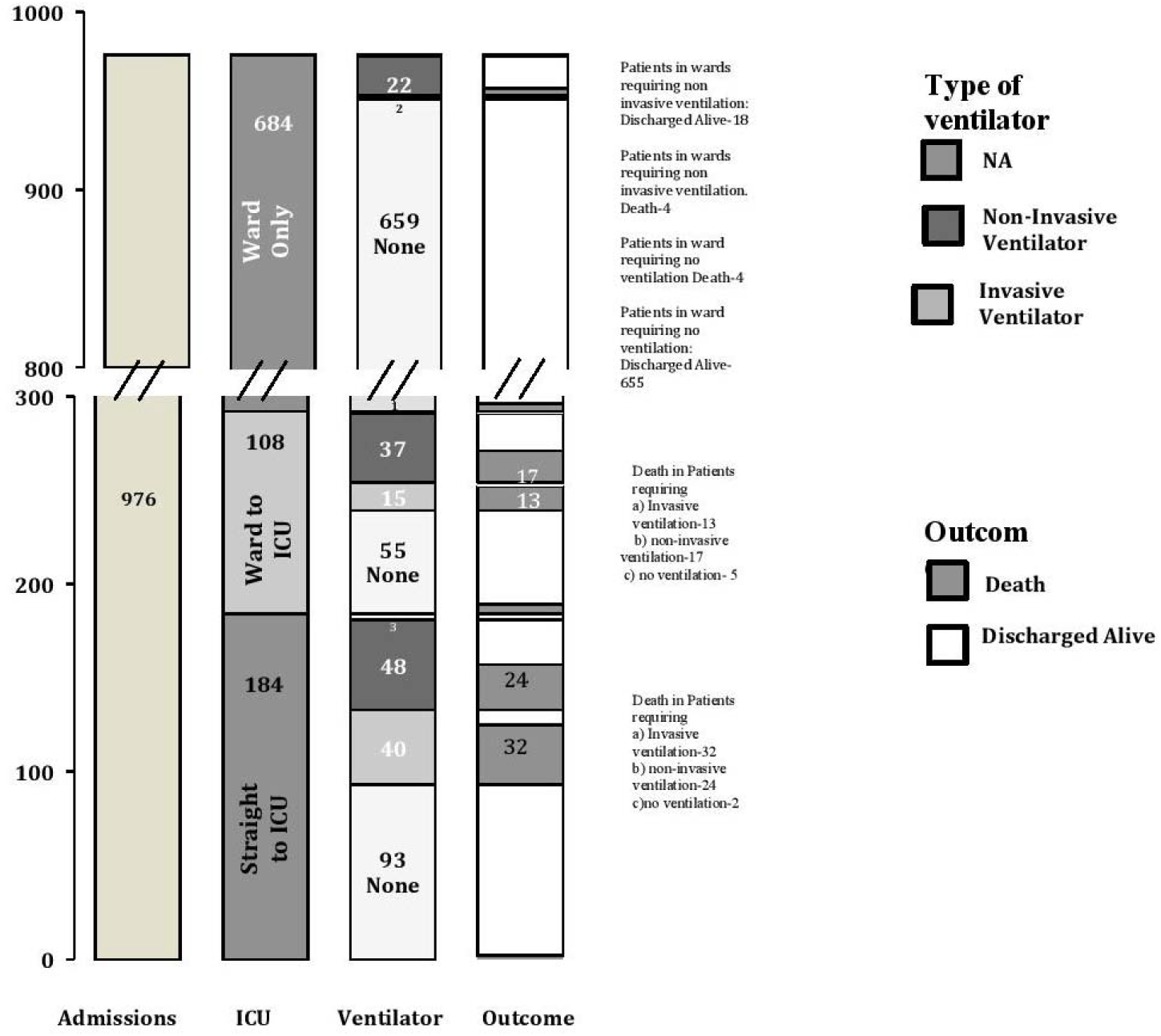
(Broken Y axis) ICU admission, ventilator requirement and outcome of the total study population. Note: 1.Patients in wards requiring ventilation are patients who came to emergency room and were placed in isolation rooms requiring invasive or non-invasive ventilation but never went to the ICU and then were shifted to the wards. 2. Missing data: We do not have the ventilator requirements for 4 patients in the ICU (3 admitted straight to ICU and 1 shifted from ward to ICU). All 4 patients were discharged alive.

Oxygen therapy was required in a total of 233 (23.87%) patients. Oxygen information in ICU was available for 288 cases out of 292, and 679 out of 684 ward cases. A total of 178 (61.81%) of 288 in ICU required oxygen therapy, whereas 55 (8.10%) out of 679 ward patients required this therapy. Mortality (33.91%) was significantly higher (P < 0.001) in those requiring oxygen therapy compared to those requiring no oxygen (3.23%). Those requiring oxygen therapy had nearly 10 times mortality.

A total of 166 (17.1%) required ventilator support – 57 invasive and 109 on non-invasive ventilator. Mortality was much higher (P < 0.001) in patients requiring ventilator support, even more so in patients requiring in invasive ventilation (82.46%) versus those requiring non-invasive ventilator (41.28%) (Table 7).

**Table 8.** Mortality in relation to medication.

| Medication | Outcome |  |  |  | Total |  |
| --- | --- | --- | --- | --- | --- | --- |
|  | Death |  | Discharged Alive |  |  |  |
|  | Count | % | Count | % | Count | % |
| All Cases | 103 | 10.55 | 873 | 89.45 | 976 | 100.00 |
| HCQ+/- Azithromycin | 69 | 8.27 | 765 | 91.73 | 834 | 85.55 |
| Ivermectin | 0 | 0.00 | 34 | 100.00 | 34 | 3.48 |
| Steroids | 57 | 47.11 | 64 | 52.89 | 121 | 12.40 |
| Convalescent Plasma Therapy | 5 | 14.29 | 30 | 85.71 | 35 | 3.58 |
| Tocilizumab | 22 | 25.00 | 66 | 75.00 | 88 | 9.01 |

Ata time when no approved treatment was available, our study observed lower mortality amongst people receiving HCQ+/- azithromycin (8.27%) compared to the overall mortality of 10.55%. Only 3.48% of the patient received Ivermectin, with 100% recovery in all cases. For steroids, the mortality is 47.11%. Similarly, for tocilizumab and convalescent plasma therapy, the mortality was 25.00% and 14.29%, respectively

### Duration of Hospital Stay

Overall median duration of stay in the hospital was 8 days (IQR: 5-11 days) and the most common (mode) was 6 days. Those in ICU stayed nearly a day longer (median 9 days) than those who never went to the ICU (median 8 days). A Mann-Whitney U test revealed that median duration of stay was significantly (P = 0.004) higher in those who were admitted to the ICU compared to those who were not.

**Table 9.** Duration of hospital stay in patients admitted to the ICU and those in the ward.

| Facility | *n* | Missing | Duration of Hospital Stay (days) |
| --- | --- | --- | --- |

|  |  |  | Mean | Median | Mode | Standard Deviation | Minimum | Maximum | Interquartile Range (days) |
| --- | --- | --- | --- | --- | --- | --- | --- | --- | --- |
| ALL | 974 | 12 | 8.77 | 8.00 | 6 | 5.13 | 0 | 65 | 5-11 |
| ICU | 290 | 02 | 9.58 | 9.00 | 6 | 6.18 | 0 | 65 | 6-12 |
| NO ICU | 674 | 10 | 8.43 | 8.00 | 5 | 4.57 | 0 | 39 | 5-10 |

## DISCUSSION

To the best of our knowledge, this is the largest study to be conducted on hospitalized patients with confirmed COVID-19 in India.

Our main findings are disproportionately higher admissions of male patients. The ones with the highest risk of mortality were elderly (60-74 years) males. The most common existing co-morbidity leading to mortality was CKD, followed by diabetes with macrovascular complications and chronic cardiac diseases. Thus, these are the patients who need to be extra careful. Nearly one-tenth had no symptoms, implying that some of those without symptoms may also require admission. Those who had usually presented with fever, productive cough, sore throat and dyspnea. Very few patients presented with gastrointestinal symptoms (11%), however this could be an under-representation since these patients fell outside the criteria for testing ^(12,13)^.

Nearly 30% required admission to ICU. The main reason for ICU admission was hypoxemia. The mortality was 32.1% compared to 1.46% in patients who did not have a ICU stay. There was no difference in mortality in those who went straight to ICU versus those shifted from ward to ICU. Thus, the prognosis of these two groups of patients was the same.

Amongst those requiring a high dependency unit, mortality was higher in those requiring some form of ventilation compared to those who did not. This information can help anticipate the clinical course that can be expected in a patient depending upon the presentation and co-morbidities. It can also help to identify those at higher risk of severe infections and mortality, and triage them accordingly.

Our study found lower mortality rates in patients receiving HCQ+/- Azithromycin. However, this can be misleading as only the mild and moderate cases received this drug. According to the treatment guidelines, HCQ was not administered in the severe cases where mortality was expected to be much higher. Thus, the lower mortality in this group is most likely due to milder disease rather than the drug itself. For ivermectin, the sample size is too small to arrive at any conclusion regarding the efficacy despite 100% recovery in the 34 patients. The mortality rate in steroid group was 47.1%, which was higher compared to that of the study population (10.55%). This high rate can be attributed to steroids being given to more severe cases at that time. However, in subsequent patients we incorporated steroids much earlier for patient care and they have shown significant mortality benefit. In the initial phase of COVID-19 in India (April-May 2020), the use of treatment modalities like steroids, tocilizumab and convalescent plasma therapy were generally restricted to late stage when the condition becomes critical, hence the mortality is much higher in these groups. Subsequent patients received these drugs much earlier in the course of the disease and have shown significant decrease in mortality.

### Comparison with Other Studies

Our patients had a lower prevalence of hypertension (23.6%), diabetes without macrovascular complications (15.3%), with macrovascular complications (9.6%) and CKD (5.4%) when compared to patients in USA, UK, Italy and China ^(14,15,16,17)^, despite these higher prevalence of this disease in the general Indian urban population as per the National Family Health Survey 2015-16. ^(18)^ This could be attributed to precautionary measures taken by the Government of India in terms of strict lockdown, ban on international and domestic travel, along with mandatory quarantine for all passengers from rescue flights before they were allowed to go home and the adherence to these rules by the general public. Being geographically closer to China may have instilled a greater fear in the people. It was surprising that not many patients admitted with COVID-19 had COPD, although this is consistent with those reported from USA and China^(19,20)^.

Overwhelming disease in developed nations like the USA, Spain and Italy possibly fostered the need to be extra cautious amongst our people. Our study subjects may represent the segment of the urban Indian population who are generally conscious about their health and have easy access to modern medical facilities. In addition, most of the doctors were available for video or telephonic consultations, minimizing the need for hospital visits. This combination of vigilance and access to healthcare from home, may have led people with co-morbidities less affected compared to some other countries.

The number of patients admitted to the ICU at our centers was higher (29.9%) when compared to USA, UK and China, however the mortality (31.8%) was less in these cases than seen in USA and UK, but more than reported from China ^(14,15,17)^. The higher admission rate to ICU could be a result of late presentation, with people trying to home quarantine in case of mild/moderate symptoms and only coming to the hospital when their condition worsened. Being tertiary care hospitals, a significant proportion of our ICU patients were from Tier – 2 & Tier – 3 cities and these patients usually came with more severe disease due to late presentation.

India was affected later than the western countries, giving us the advantage of learning how other countries dealt with the pandemic. This gave us ample time to improvise our infrastructure and adapt to the inevitable need of increased ICU beds, ventilators, and PPE kits. Also, we had the advantage of well-defined guidelines created both at the central level by the government and at the respective hospital level. A large number of therapies such as use of convalescent sera, anti-viral drugs like remdesivir ^(21,22,23)^ and antibody against IL-6 receptors (Tocilizumab ^(24,25)^) were already tried and proven to be efficacious in multiple centers across the globe. Since this data is only for the first 1000 patients, remdesivir was not used due to unavailability. Out of the total 6000 patients treated at our center thus far (by September 2020) many received remdesivir as soon as it became available in the country.

Tocilizumab was initiated and 9% of the patients received them in this series. Due to high demand, there were shortages in the drug supply in the initial period, but now it is widely available and all the patients requiring it have been administered the drug henceforth.

One of our centres was among the first in the country to provide plasma therapy through a randomized controlled trial, and though these numbers were very few in the first 1000 patients, it showed better outcome amongst some sections of the patients. Convalescent plasma therapy has been administered to over 500 patients subsequently.

Amongst the patients admitted to the ICU, the requirement of ventilation in our cases (48.2%) was much lower than in USA (85%) and Italy (71%). ^(16,27)^

The overall mortality in our inpatient hospital cohort was 10.5%, which was also much lower than those reported for the USA (21.1%) and UK (26%) at that time ^(14,15)^This favorable outcome again may be ascribed to the ability of our healthcare system to adapt and ensure availability of resources. The recovery rate in India was 73.5% at the time of this report compared to 53.2% in the USA and 74.9% in Brazil^(28)^. However, the recovery in Delhi was high at 90%, pointing to availability of healthcare resources – both personnel and infrastructure. However, if the number of cases increases exponentially as feared, there might be a shortage of beds, ventilators, and other vital resources. Our guidelines are constantly updated to ensure best possible care for patients in an evolving situation.

The overall median duration of stay in the hospital both for ward and ICU was considerably less in our patients when compared to US (9 days in discharged and 28.5 in still admitted) and China (12 days) ^(14,17)^. This can be attributed both to a younger population in our cohort and early aggressive treatment that obviated the need for ICU admission and eventually mechanical ventilation. The reason for the low absolute difference between the median duration of stay seen between the ward patients (8 days) and the ICU patients (9 days) was due to government guidelines of continued admission until the patient tested negative twice. Beginning May, these guidelines allowed discharge after 3 days of being afebrile without being retested. Currently, our facility has a much lower duration of stay amongst ward patients in comparison to ICU patients.

### Implications for Clinical Practice

A pandemic is a time when every country is bound to face shortage of resources. Our study provided evidence that elderly males and people with co-morbidities to have significantly higher ICU admission rates, ventilator requirement and mortality rates. With the rising numbers and evident need for cautious resource allocation, this information can help us triage patients; ensure timely hospitalization and a watchful eye on these high-risk groups. We also noticed fewer deaths in our ICU as compared to other countries owing to early aggressive treatments and resource availability (ventilator). This helped us to ensure the appropriate use of treatment modalities like steroids, tocilizumab, and plasma therapy in patients with increasing need for oxygen therapy. This clinical management strategy reduced the mortality rate even further in subsequent patients. This also reduced ICU admission and should therefore be used early on especially in the high-risk group and people with dyspnea and fever. Our experience suggests that every institution should create a standard protocol incorporating latest evidence-based treatment and should be followed for every patient. We are now analyzing the next 5000 COVID positive admissions depending upon the type of treatment they were given and the outcome thereafter, and the time from increased oxygen need to aggressive treatment initiation that will probably give us more detailed information on the use of different pharmacomodalities and when to start them.

## LIMITATIONS

Not all of our centers have Electronic Medical Records and tracking of written records was challenging and led to some missing information. We removed 24 patients for whom a lot of information was missing, however, even in the 976, there were a few missing entities. Secondly, our patients had limited access at that time to therapies like convalescent plasma due to the challenge of securing voluntary plasma donation and short supply of drugs like tocilizumab and remdesivir (which became available in late June). No follow up was done post discharge to check for any post COVID illness, signs or resurgence of symptoms in this cohort; however, we are now conducting telephonic follow up on our patients for upto 6 weeks post discharge with a plan to further study recovery post discharge. We had insufficient data regarding weight and height hence body mass index (BMI) and obesity, which have shown strong association with COVID-19, could not be studied in our patients. Our study subjects are mostly drawn from an urban setting and the findings might be specific to urban areas only. COVID 19 is entirely a new disease that has become rampant worldwide, and we may have missed unforeseen factors.

## CONCLUSION

The study observed that the elderly male population is at considerably higher risk of mortality from COVID-19. Also, the patients with co-morbidities have increased chances of worse outcomes. These are powerful predictors for requirement of hospital admission in these patients instead of outpatient care. We strongly believe that appropriate triaging of patients depending upon age, presence of co-morbidities and severity of disease followed by early institution of treatment as per the standard guidelines, adequately trained man force, adequate PPE supply and good critical care services can go a long way in bringing down the mortality in this disease.

## Data Availability

The study is based on institutional records.

## Acknowledgement

We thank Dr. Menka Loomba, Ms. Archa Misra, Ms. Akanksha Tyagi, Ms. Shruti Singh for their technical support. We deeply appreciate the dedication and effort of our frontline healthcare workers who have been working round the clock to help us tide over this crisis. Finally, we express our heartfelt despair for the loss of our patients and their families.

## Ethical Approval

This study was approved by the ethics committee of MaxHealthcare (RS/MSSH/DDF/SKT-2/IEC/IM/20-16)

## Author contributions

Study conception and design: SB,AS. Acquisition, analysis, or interpretation of data: AI, VJ, AD, OS, YPS, IC, VA,RP,AA, AS. Drafting of the manuscript: AS. Critical revision of the manuscript for important intellectual content: SB, AI, VJ, SJ. Statistical analysis and interpretation: AI. Administrative, technical, or material support: VJ, AD, OS, YPS, IC, VA, RP, AA. Study supervision: SB, SJ, VJ. The corresponding author attests that all listed authors meet authorship criteria and that no others meeting the criteria have been omitted.

